# Deep learning-based detection of COVID-19 using wearables data

**DOI:** 10.1101/2021.01.08.21249474

**Authors:** Gireesh K. Bogu, Michael P. Snyder

**Affiliations:** Department of Genetics, Stanford University School of Medicine, Stanford, CA, USA

## Abstract

**Background:** COVID-19 is an infectious disease caused by SARS-CoV-2 that is primarily diagnosed using laboratory tests, which are frequently not administered until after symptom onset. However, SARS-CoV-2 is contagious multiple days before symptom onset and diagnosis, thus enhancing its transmission through the population.

**Methods:** In this retrospective study, we collected 15 seconds to one-minute heart rate and steps interval data from Fitbit devices during the COVID-19 period (February 2020 until June 2020). Resting heart rate was computed by selecting the heart rate intervals where steps were zero for 12 minutes ahead of an interrogated time point. Data for each participant was divided into train or baseline by taking the days before the non-infectious period and test data by taking the days during the COVID-19 infectious period. Data augmentation was used to increase the size of the training days. Here, we developed a deep learning approach based on a Long Short-Term Memory Networks-based autoencoder, called LAAD, to predict COVID-19 infection by detecting abnormal resting heart rate in test data relative to the user’s baseline.

**Findings:** We detected an abnormal resting heart rate during the period of viral infection (7 days before the symptom onset and 21 days after) in 92% (23 out of 25 cases) of patients with laboratory-confirmed COVID-19. In 56% (14) of cases, LAAD detection identified cases in their pre-symptomatic phase whereas 36% (9 cases) were detected after the onset of symptoms with an average precision score of 0·91 (SD 0·13, 95% CI 0·854–0·967), a recall score of 0·36 (0·295, 0·232–0·487), and a F-beta score of 0·79 (0·226, 0·693–0·888). In COVID-19 positive patients, abnormal RHR patterns start 5 days before symptom onset (6·9 days in pre-symptomatic cases and 1·9 days later in post-symptomatic cases). COVID-19+ patients have longer abnormal resting heart rate periods (89 hours or 3·7 days) as compared to healthy individuals (25 hours or 1·1 days).

**Interpretation:** These findings show that deep learning neural networks and wearables data are an effective method for the early detection of COVID-19 infection. Additional validation data will help guide the use of this and similar techniques in real-world infection surveillance and isolation policies to reduce transmission and end the pandemic.

**Funding:** This work was supported by NIH grants and gifts from the Flu Lab, as well as departmental funding from the Stanford Genetics department. The Google Cloud Platform costs were covered by Google for Education academic research and COVID-19 grant awards.

**Research in context:** *Evidence before the study:* COVID-19 resulted in up to 1·7 million deaths worldwide in 2020. COVID-19 detection using laboratory tests is usually performed after symptom onset. This delay can allow the spread of viral infection and can cause outbreaks. We searched PubMed, Google, and Google Scholar for research articles published in English up to Dec 1, 2020, using common search terms including “COVID-19 and wearables”, “Resting heart rate and viral infection”, “Resting heart rate and COVID-19”, “machine learning and COVID-19” and “deep-learning and COVID-19”. Previous studies have attempted to use an elevated resting heart rate as an indicator of viral infection. Although these studies have investigated the early prediction of COVID-19 using resting heart rate and other wearables data, studies to investigate a deep learning-based prediction model with performance evaluation metrics at the user level has not been reported.

*Added value of this study:* In this study, we created a deep-learning system that used wearables data such as abnormal resting heart rate to predict COVID-19 before the symptom onset. The deep-learning system was created using retrospective time-series datasets collected from 25 COVID-19+ patients, 11 non-COVID-19, and 70 healthy individuals. To our knowledge, this is the first deep-learning model to identify an early viral infection using wearables data at the user level. This study also greatly extends our previous phase-1 study and factors unpredictable behavior and time-series nature of the data, limited data size, and lack of data labels to evaluate performance metrics. The use of a real-time version of this model using more data along with user feedback may help to scale early detection as the number of patients with COVID-19 continues to grow.

*Implications of all the available evidence:* In the future, wearable devices may provide high-resolution sleep, temperature, saturated oxygen, respiration rate, and electrocardiogram, which could be used to further characterize an individual’s baseline and improve the deep-learning model performance for infectious disease detection. Using multi-sensor data with a real-time deep-learning model has the potential to alert individuals of illness prior to symptom onset and may greatly reduce the viral spread.

## Introduction

COVID-19 is a contagious respiratory disease caused by the novel coronavirus, severe acute respiratory syndrome coronavirus 2 (SARS-CoV-2).^1^ As of 6 December 2020, more than 66·7 million people have been infected world-wide with 14·6 million in the U.S alone, and the number of cases continues to rise.^4^ COVID-19 PCR and antigen testing, shelter-in-place, and social distancing are effective and complementary public health strategies, but in their patchwork implementation, have not been adequate to stop ongoing virus transmission. Viral load (number of viral particles) and duration of viral shedding (where the virus replicates and is released into the environment by an asymptomatic or symptomatic host) are important determinants for COVID-19 transmission.^2,3^ Based on current literature, viral shedding of SARS-CoV-2 may begin 5 to 6 days before the onset of symptoms and then declines 14·6 days to 17·2 days following symptom onset, although in some cases of protracted infection, shedding may continue for much longer.^2,3^ In contrast to the etiology of related coronaviruses, such as SARS-CoV-1, where symptom onset was rapid and enabled swift isolation procedures, the long asymptomatic contagious period of COVID-19 adds urgency to the issue of early infection detection to guide testing, treatment, and isolation policies and reduce spread.

Tracking biometric data from wearable devices is a promising method for detecting COVID-19^5–9^ and other respiratory viral infections.^10^ Wearable devices contain a myriad of different sensors that collect distinct data types such as heart rate, steps, and sleep, which can potentially be used to track viral infections over time and proactively detect their onset using statistical methods like cumulative sum (CUSUM), RHR-Diff, and HROS-AD, as in our previous work.^5^ However, physiological measurements are often affected by external factors or variables such as environmental conditions (high temperature and altitude etc.), and this can lead to inherently unpredictable time-series data.^11^ Detecting anomalies in these scenarios can be challenging with standard approaches based on statistical measures that use static data or a pre-specified time-window to detect changes in the underlying distribution or prediction errors.^11^

Here, we employed a deep learning framework, Long Short Term Memory Networks (LSTM)-based Autoencoder for Anomaly Detection (LAAD), that learns temporal dependencies from the input data (baseline or training data) to reconstruct ‘normal’ time-series output, calculate a threshold based on the reconstruction error from the baseline, and uses it to detect anomalies in the test data.^11^ LAAD contains an encoder that learns a vector representation of the input time-series data and a decoder that uses this representation to reconstruct the baseline data. For the implementation of both the encoder and the decoder, we use LSTM cells. LSTMs can learn dependencies from one instance to another while solving the vanishing-gradient problem that affects standard recurrent neural networks.^11^ This capability is due to a more complex cell architecture that accurately maintains a memory of important correlations. LAAD’s ability to learn higher-level temporal patterns without prior knowledge of the pattern duration and its ability to learn from ‘normal’ data makes it robust to both predictable and unpredictable time-series data.^11^

## Methods

### Datasets

We used publicly shared data from a previously-published Phase-1 study.^5^ We selected 25 COVID-positive, 11 non-COVID-19 illness, and 70 healthy individuals with enough data to train, test and evaluate metrics. It also contains metadata with self-reported symptom dates and diagnosis dates. We used a *sample* function from *pandas* library (https://pandas.pydata.org) to randomly assign a symptom date for each healthy individual using their test data time intervals with a fixed random seed. For two COVID positive datasets without symptom dates, we used diagnosis dates as symptom dates.

### LAAD framework

#### Pre-processing

For each user, we selected the heart rate data and merged the step data with the same exact time-stamp as heart rate. Merged data was aggregated to one-minute resolution using the *resample* function from *pandas* library. Next, the resting heart rate was calculated by selecting the heart rate intervals where steps were zero for 12 minutes ahead of a given time point. Finally, we used moving-averages (mean = 400 hours) to smooth the time-series data with the *rolling* function from *pandas* and further aggregated to one-hour resolution by taking the mean.

#### Data splitting and normalization

We labeled the data as infectious, non-infectious and recovery periods based on general consensus of the recent COVID-19 literature.^1–3^ Seven days before and 21 days after the symptom onset were considered as infectious, 10 to 20 days before the infectious period as non-infectious period and days after infectious period as a recovery period (**figure 1A**). Using these labels we divided the data into training data or baseline using the days before the non-infectious period and test data by taking the days during the non-infectious, infectious period and recovery periods.

**Figure 1.**
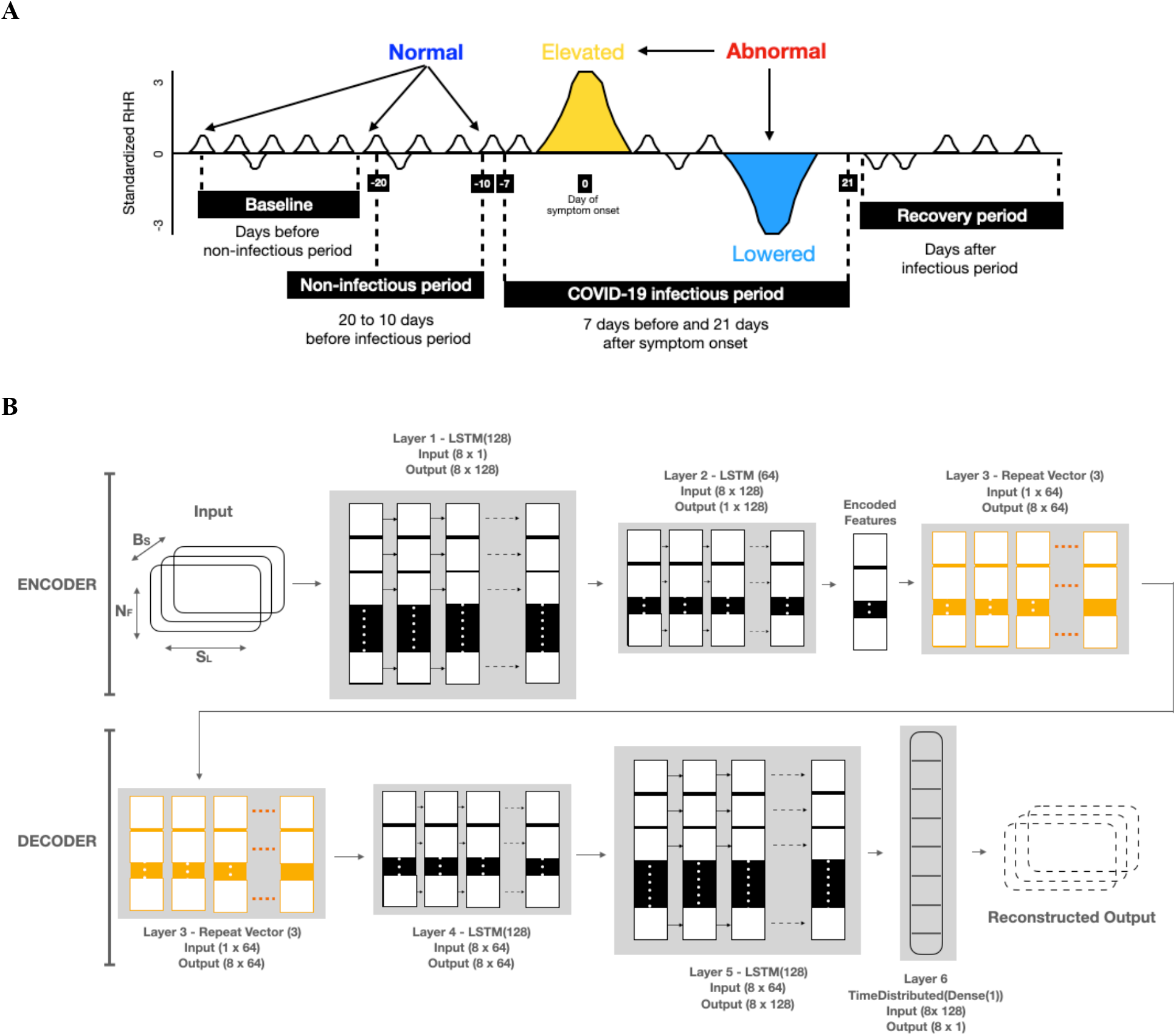
Data design and LAAD framework (LSTM Encoder-Decoder inference steps for input to reconstruct the output). (**A)** To detect abnormal RHR (resting heart rate) data was split into baseline (train) and test sign symptom onset day as a reference. First, test data was set by taking 20 days prior to symptom onset and 21 days after. Second, test data was split into infectious period (7 days prior symptom onset and 21 days after) and non-infectious period (20 to 10 days before infectious period), and recovery period (days after infectious period). If the RHR during the infectious period is changed (elevated or lowered) from its user’s baseline, it would be classified as “abnormal RHR”. Further, to evaluate the model performance, predictions in the infectious period were compared against the non-infectious period. (**B**) LAAD takes baseline standardized RHR data of shape (B_S_, S_L_, N_F_), where B_S_ is batch size, N_F_ is the number of features and S_L_ is sequence length or time steps and passes it to the first layer. The input data has 8 timesteps and one feature. First layer has as many LSTM cells as the S_L_ and makes each cell per timestep emit a signal to a second layer. Layer 1, LSTM(128), reads the input data and outputs 128 features with 8 timesteps. Second layer has half the size of LSTM cells than the previous and only the last cell emits an output. Layer 2, LSTM(64), takes the 8×128 input from Layer 1 and reduces the feature size to 64. The output of this layer is the encoded feature vector of the input data. Third layer uses a Repeat Vector that replicates the feature vector 3 times and gets a 2D array for the fourth layer (1st LSTM layer in Decoder) and acts as a bridge between encoder and decoder. The decoder layers unfold the encoding by stacking LSTM layers in the. reverse order of the encoder. Layer 4, LSTM (64), and Layer 5, LSTM (128), are the mirror images of Layer 2 and Layer 1, respectively. Layer 6, TimeDistributed (Dense(1)), is added in the end to get the reconstructed output, where “1” is the number of features in the input data.

Training data is further divided into 95% training and 5% validation data after applying data augmentation (**Methods**). Train data and test data were standardized separately and transformed into a tensor format before feeding them into the LAAD framework. Data was standardized by setting the mean to zero and variance to one with the help of a standard *scaler* from s*klearn* library (https://scikit-learn.org).

#### Data windowing

We split the RHR sequence and grouped the resulting data using a fixed-length window (W) of size 8. The value of W defines how many time-lags are processed by the LAAD that classifies the input as an anomaly or not. Finally, we reshaped the data format to tensor with features: number of samples (batch size), number of time steps per sample (sequence length), number of features.

#### Data augmentation

We applied seven time-series data augmentation techniques found in the literature.^12,13^ *Scaling*-changes the magnitude of the data in a window by multiplying by a scalar. The scalar was determined by a Gaussian distribution with mean 1 and standard deviation 0·1. *Rotations*-applies arbitrary rotations to the existing data. *Permutation*-randomly perturbs the temporal location of within-window events. To perturb the location of the data in a single window, we first slice the data into N same length segments, with N ranging from 1 to 5, and randomly permute the segments to create a new window. *Magnitude-warping* – changes the magnitude of each time series is multiplied by a curve created by a cubic spline with four knots at random magnitudes with a mean 1 and standard deviation 0·2. *Time-warping*-perturb the temporal location using a random smooth warping curve generated by a cubic spline with four knots at random magnitudes with a mean 1 and standard deviation 0·2. *Window-warping* - selects a random window of 10% of the original data and either speed it up by 2 or slows it down by 0:5. *Window-slicing* - a window of 90% of the original time series is chosen at random. We created eight times the size of the baseline data by applying scaling on the baseline data and rotations on scaled data and so on.

#### LSTM-based Autoencoder

We used a sequence-to-sequence autoencoder since our data consists of time-series sequences (**figure 1B**). The objective was to reconstruct the RHR data using an encoded representation of the input (baseline) time-series sequences. The autoencoder consists of an encoder and decoder.^11^ An LSTM encoder learns a fixed-length vector representation of the input RHR time-series data and the LSTM decoder uses this representation to reconstruct the RHR time-series using the current hidden state and the value predicted from the previous time-step. For the implementation of both the encoder and the decoder, we used LSTM layers, which consider temporal dependencies from one sequence to another. We trained the autoencoder using the baseline data with baseline RHR temporal sequences and reconstructed it with a low reconstruction error and then used it to detect anomalies in test data.

We used *Talos* (https://github.com/autonomio/talos) to evaluate the algorithm performance by measuring the reconstruction error with a different set of hyper-parameters and considered the model that gave the lowest error as best. Early stopping callback was used to avoid overfitting by stopping the training at the optimal time.

For LAAD, we used 4 LSTM layers: one RepeatVector layer, one TimeDense layer, and 128 hidden neurons for implementing both the encoder and decoder. The reconstruction error is calculated as an MSE (Mean Squared Error) and the ADAM algorithm is used to optimize the learning process. We split the training dataset and used 5% of that data as a validation set for evaluation and monitoring validation loss. We set a value (maximum value) of reconstruction error from the baseline data as a threshold and annotated any value in the test data that is greater than this threshold as an anomaly.

### Anomaly distance, signal strength

Anomaly distance was calculated by subtracting the date and time of the first anomalous event during the COVID-19 infectious period from the symptom onset date. Anomaly signal strength was calculated by dividing the number of anomalous events in a pre-symptomatic window (7 days before the symptom onset) with the loss calculated by mean square error (MSE) and further divided by the length of the window (7 days) and multiplied by 100. In post-symptomatic cases, 21 days after the symptom onset and 21 days window length were used. Cases that have more than 6 anomaly signal strength scores were grouped as strong and those with less than 6 were grouped as weak.

### Number of abnormal RHR hours, delta abnormal RHR

For each user, a number of abnormal RHR hours were counted during COVID-19 infectious period. For each user, delta RHR was calculated by subtracting the total RHR of the anomalies in test data (COVID-19 infectious period) from baseline/training data. Delta RHR was further grouped as elevated if the RHR was positive and lowered if the RHR was negative.

### Performance evaluation

True positives (TP) are the number of anomalous RHR hours that are correctly identified as anomalous. False positives (FP) are the number of normal hours that are incorrectly identified as anomalous. True negatives (TN) are the number of normal hours that are correctly identified as normal. False negatives (FN) are the number of anomalous hours that are incorrectly identified as normal. We calculated the performance metrics as follows: precision-recall, where precision is defined as the ratio between true positives and the sum between true positives and false positives (Precision = TP / (TP + FP)) and recall (also known as sensitivity or true-positive rate) is defined as the ratio between the true positives and the sum between true positives and false negatives (Recall = TP / (TP + FN)). Precision measures the proportion of anomalous hours that are relevant and recall measures of how many hours are anomalous. Furthermore, we calculated the F-beta score, a weighted mean of both precision and recall. The F-beta score is a generalization of the F-score that adds a configuration parameter called beta. A default beta value is 1·0, which is the same as the F-score. We used a beta value, such as 0·1, that gives more weight to precision and less to recall, assuming false positives are more important to minimize, but false negatives are still important (F-beta = ((1 + beta^2^) x Precision x Recall) / (beta^2^ x Precision + Recall)).

### Visualization

All the plots were generated using the following libraries - Matplotlib https://matplotlib.org/; Seaborn https://seaborn.pydata.org/; ggplot https://ggplot2.tidyverse.org/.

## Results

We collected heart rate and steps wearable data from 106 individuals, including 25 COVID-19+, 11 non-COVID-19 illness with self-reported symptoms, and 70 healthy individuals with no symptoms or illness (**table 1; appendix 2 pp 1**) during the COVID-19 period (February 2020 until June 2020) to examine whether there is any consistent anomalous signal during the COVID-19 infectious period (defined as 7 days prior and 21 days after symptom onset) (**figure 1A**). Before applying the deep learning framework, we inferred resting heart rate (RHR) from heart rate and steps data and then aggregated resulting RHR data to one-hour resolution. We defined the anomalous signal or abnormal RHR based on its deviations (either elevated or lowered) from the baseline (**figure 1A**). We then divided the wearable data into baseline or training data using the days before the non-infectious period and test data using the days during the non-infectious and the infectious period (**figure 1A; appendix 2 pp 2)**. Next, we transformed univariate RHR data into sub-sequences by combining time steps contiguous data values using a data windowing method.

**Table 1.** Data summary. In total, 106 datasets and 3 groups. Data was split between training and test (Test_n_ – test-normal or data during the non-infectious period, Test_a_ – test-anomaly or data during the infectious period) using symptom onset day as a reference and the values are shown were the average number of hours (standard deviation) per group.

| Group | Number of individuals | Train | Test | Test<sub>n</sub> | Test<sub>a</sub> |
| --- | --- | --- | --- | --- | --- |
| COVID-19-positive | 25 | 333 (233) | 713 (350) | 124 (52.1) | 336 (159) |
| Non-COVID-19 | 11 | 266 (171) | 578 (399) | 105 (53.1) | 273 (152) |
| Healthy | 70 | 211 (81.9) | 575 (340) | 81 (57.4) | 235 (137) |

Deep learning models typically require large datasets for training. Otherwise, the model is prone to overfitting. In our case, we only have a limited number of days available for training (**appendix 2 pp 2**). Augmented data can cover unexplored input space, prevent overfitting, and improve the generalization ability of a deep learning model.^12,13^ To do this, we applied different data augmentation techniques^12,13^ on our limited training data by transforming each user’s training data into approximately eight times the size.

Next, we built an LAAD model that takes input with a specific shape (batch size, sequence length, number of features) and learns the structure by reconstructing an output of the same shape (**figure 1B**). We used augmented training data as the input to the LAAD model. We further split training data into 95% training and 5% validation sets and used these to train the LAAD model. To get optimal performance, we used tuned hyper-parameters (hidden layers = 6, number of neurons = 128, batch size = 64, number of epochs = 1200 with early stopping callbacks, learning rate = 0·0001, optimizer = adam). Using a reconstruction loss technique, mean square error (MSE), we built a threshold where, if the MSE for a test sample is greater than this threshold, that sample is labeled as an anomaly or abnormal RHR (**appendix 1 pp 4**,**5; appendix 2 pp 3-5**).

Using LAAD and data from the COVID-19 patients, we detected an abnormal RHR signal in 14 individuals during pre-symptomatic (early), in 9 individuals during post-symptomatic periods (late), and failed to detect a signal in two individuals (missed) (**figure 2A; appendix 1 pp 6, appendix 2 pp 6**). As an example, in the COVID-19 positive individual, ASFODQR, the anomaly score representing the level of abnormal RHR starts before symptom onset and increases during the COVID-19 infectious period as compared to the baseline, non-infectious or recovery periods (**figure 2B**). Abnormal RHR starts 5·04 days before symptom onset, comprising 6·94 days early in pre-symptomatic cases and 1·92 days late in the post-symptomatic cases (**figure 2C; appendix 2 pp 6**). Of 14 pre-symptomatic cases, 7 had strong and 7 had weak abnormal RHR signals during the pre-symptomatic period, and of the 9 post-symptomatic cases, 4 had strong and 5 had weak abnormal RHR signals during the post-symptomatic period (**figure2A**; **appendix 2 pp 6**). This indicates that not all COVID-19 positive individuals have similar levels of abnormal RHR and also suggests that LAAD can successfully detect cases where the signal is not strong.

**Figure 2.**
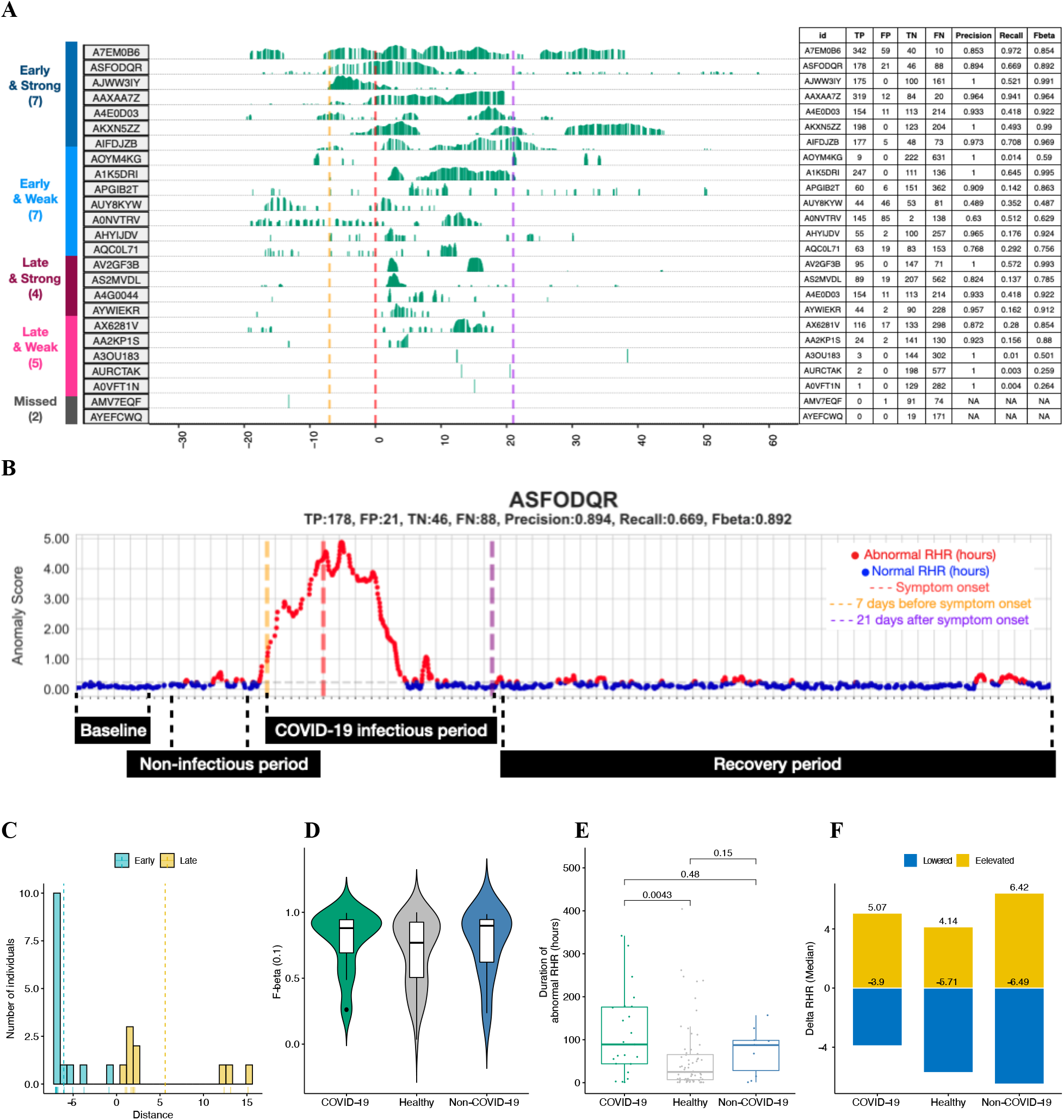
LAAD predictions, Summary of detection timing, Evaluation metrics, Comparison of abnormal RHR between COVID-19, healthy and non-COVID-19 groups. (**A**) On the left, bar plots showing anomaly scores obtained from reconstruction error or mean squared error (MSE) and timings of infection detection from the LAAD model with respect to the different periods of SARS-CoV-2 infection for COVID-19+ participants. Based on the time of detection, participants were grouped into early and late groups. Based on anomaly signal strength, these groups were further sub-grouped into strong and weak. Participants were ordered based on early and strong to weak and late grouping order. One participant, whose LAAD failed to detect anomaly during infectious period was annotated as “missed”. The x-axis shows days before, during and after the infection and the y axis shows loss values from LAAD (green bars). On the right, a table showing performance metrics (TP - true positives, FP – false positives, TN – true negatives, FN - false negatives, precision, recall and F-beta score) for each participant. The self-reported symptom onset date was shown as a red dotted line, a pre-symptomatic window as a gold dotted line and a post-symptomatic window as a purple dotted line. (**B**) Scatter plot showing the distribution of anomaly score (MSE loss) of COVID-19+ individual (ASFODQR) highlighting the baseline (first 10 days), non-infectious (10 days prior to the infectious period, −20 to −10), infectious (−7 to −21) and recovery periods (days after the infectious period). Abnormal RHR was shown in red and normal RHR in blue color. Self-reported symptom onset date was shown as a red dotted line, a pre-symptomatic window as a gold dotted line and a post-symptomatic window as a purple dotted line, and anomaly threshold was shown as a horizontal grey dotted line. (**C)** Bar plot showing the distribution of detection timing during the infectious period (blue = early or pre-symptomatic, gold = late or post-symptomatic) in COVID-19+ patients. (**D)** Bar plots showing the number of individuals who had more than 3 days of abnormal RHR signals during the infectious period (green = COVID-19, grey = healthy, blue = non-COVID-19). (**E**) Boxplots showing the distribution of the number of abnormal hours during infectious period in COVID-19, healthy and non-COVID-19 groups. P-values were shown on top of the boxplot. A Wilcoxon test was used to calculate the p-value between the groups. (**F)**, Bar plots showing delta RHR during infectious period in COVID-19, healthy and non-COVID-19 groups. Delta RHR of elevated RHR regions were shown in gold and lowered RHR regions shown in blue.

To measure the performance of this model, we further divided test data into a) test-normal by analyzing the days during the non-infectious period and b) test-anomaly by examining the infectious period, and evaluated predictions against each other (**figure 1A; table 1; appendix 2 pp 2**). On average, the anomaly detection model had a precision of 0·91 (SD 0·13, 95% CI 0·854–0·967), a recall of 0·36 (0·295, 0·232– 0·487), and a F-beta score of 0·79 (0·226, 0·693–0·888) (**appendix 2 pp 9**). Precision, P, was defined as the ratio between true positives, TP (the number of samples or RHR hours belonging to the anomaly class that are correctly classified as anomalous) and the sum of TP and false positives, FP, where FP represents the number of samples belonging to the normal class that are incorrectly classified as anomalous. Recall, R, was defined as the ratio between the TP and the sum of TPs and false negatives FN, which are the number of samples belonging to the anomaly class that are incorrectly classified as normal. Finally, the F-beta score is defined as the weighted mean of P and R. This allows a model to be evaluated by using a single score that accounts for both precision and recall performance.

Next, we investigated 11 non-COVID-19 individuals and found abnormal RHR signals in seven individuals during the pre-symptomatic period and two individuals during the post-symptomatic period but failed to detect a signal in two individuals (**appendix 1 pp 1; appendix 2 pp 4**,**7**). On average, the anomaly detection model had a precision of 0·803 (0·262, 0·602–1), a recall of 0·472 (0·405, 0·16– 0·783), and an F-beta score of 0·771 (0·252, 0·577–0·965) (**appendix 2 pp 10**). We further investigated 70 healthy individuals by randomly assigning a symptom date for the test data and detected abnormal RHR signal in 44 individuals during the randomly selected pre-symptomatic period and 15 individuals during the randomly selected post-symptomatic period and failed to detect a signal in 11 individuals (**appendix 1 pp 2; appendix 2 pp 5**,**8**). On average, the anomaly detection model had a precision of 0·803 (0·262, 0·726–0·88), a recall of 0·289 (0·336, 0·19–0·387), and a F-beta score of 0·703 (0·259, 0·627–0·78) (**appendix 2 pp 9-11**). Overall, LAAD performed well across all three groups with an average F-beta score of 0·75 (SD 0.04) (**figure 2D**).

COVID-19+ cases showed longer hours of abnormal RHR (89 hours or 3·7 days) during the infectious period as compared to non-COVID-19 (87·5 hours, 3·65 days) and healthy (25 hours, 1·04 days) cases, suggesting that COVID-19 infection lasts longer than anomalous periods for healthy cases and slightly longer than other types of infections (**figure 2E; appendix 1 pp 3; appendix 2 pp 12**). In total, 78·3% (18 of 23 cases) of COVID-19+ cases had more than one day of abnormal RHR signal compared to 70% (7 of 10 cases) of non-COVID19 and 51·56% (33 of 64 cases) of healthy cases (**appendix 1 pp 3**). These results suggest that the duration of abnormal RHR can predict COVID-19 and discriminate from healthy cases but not from non-COVID-19 cases. Abnormal RHR levels during the infectious period were elevated by 5·1 days and lowered by 3·9 days compared to the baseline (Delta RHR) (**figure 2F; appendix 1 pp 3; appendix 2 pp 13**). In non-COVID-19 cases and healthy individuals, abnormal RHR was elevated for 4·1 days and 6·4 days and lowered by 5·7 days and 6·5 days, respectively (**figure 2F; appendix 2 pp 13**). These results suggest that the levels of abnormal RHR can predict COVID-19 but cannot discriminate from non-COVID-19 cases and healthy cases.

## Discussion

We demonstrate that deep learning can identify periods of COVID-19 and is more sensitive than approaches used in other previously reported studies.^5,10^ However, our study has several limitations. First, all symptom onset dates were self-reported by the patients, usually after diagnostic confirmation of COVID-19. Since the data was divided into training and test sets and all metrics calculations were based on the self-reported symptom onset date, errors in self-reporting can introduce bias in our results and model performance. Second, we divided the symptomatic period into pre-symptomatic and post-symptomatic periods using recent studies on the viral infectiousness period. However, the length of these periods could vary substantially from one patient to another and thus introduce bias in our results. Third, none of the healthy patients had COVID-19 tests, and it is therefore possible that some of them had asymptomatic infections. Indeed, we found 11 healthy cases where abnormal RHR was detected for 3·7 days (89 hours) more during the infectious period, similar to COVID-19 patients (**appendix 2 pp 12**).

Fourth, on average only approximately 3 months of data was collected per user and thus deriving training, validation and test data from such limited data may be a limiting factor for the model performance. Fifth, we did not test any confirmed COVID19 negative cases, which limits the potential of our study. Sixth, only 25 COVID-19 cases were used in the analysis. Adding more samples will improve our understanding of wearable data performance in detecting COVID-19. Seventh, all data used in this study were collected from Fitbit smartwatch heart rate and activity data. Adding other data types like temperature, saturated oxygen, sleep, and data from other devices such as the Apple watch or Garmin watch could potentially improve COVID-19 detection. Eighth, choosing a correct baseline with enough data size to train the model has a major impact on predictions. For example, A7EM0B6 with only 4 days of training data showed severe underfitting of the model (**appendix 2 pp 4**). Ninth, sleep data was inconsistent and not the same resolution as heart rate and activity data, which limited the inclusion of sleep data in our model. Tenth, no cross-validation was performed to test the performance of the deep learning model on unseen datasets. Testing new COVID-19+ patients data with LAAD could validate and potentially improve the model. Finally, this deep learning framework uses retrospective data and is designed as a proof-of-concept. It has yet to be tested in a prospective real-time fashion using pre-annotated labels from users.

Overall, our work suggests that wearable sensor data could be used as a marker for early prediction of COVID-19. This study also greatly extends our previous phase-1 study^5^ and factors unpredictable behavior and time-series nature of the data, limited data size, and lack of data labels to evaluate performance metrics. A detailed real-time wearable study on the COVID-19 patients with symptoms annotated by users and confirmed by laboratory tests will further our understanding about tracking, modeling, and detecting outbreaks of SARS-CoV-2.

## Data Availability

De-identified data is available at https://storage.googleapis.com/gbsc-gcp-project-ipop_public/COVID-19/COVID-19-Wearables.zip and code is open-sourced for non-commercial purposes, and available at https://github.com/gireeshkbogu/LAAD.

## Contributors

GKB conceived the deep-learning study design and did the data analysis, data interpretation, figure creation, and writing of the manuscript. MPS revised the manuscript and provided scientific input.

## Acknowledgments

We thank D. Berrent from Survivor Corps for assistance with recruitment, and A. McDonough and T. Helgren from Fitbit Inc. for help with accessing the Fitbit data. We thank Fitbit Inc. for promoting this study and for the donation of devices. The Stanford Healthcare Innovation Lab gratefully acknowledges the support of A. Duisberg.

## Supplemental Materials

### Supplemental appendix 1

**Section 1: LAAD predictions for the non-COVID-19 group**.

**Figure S.1.**
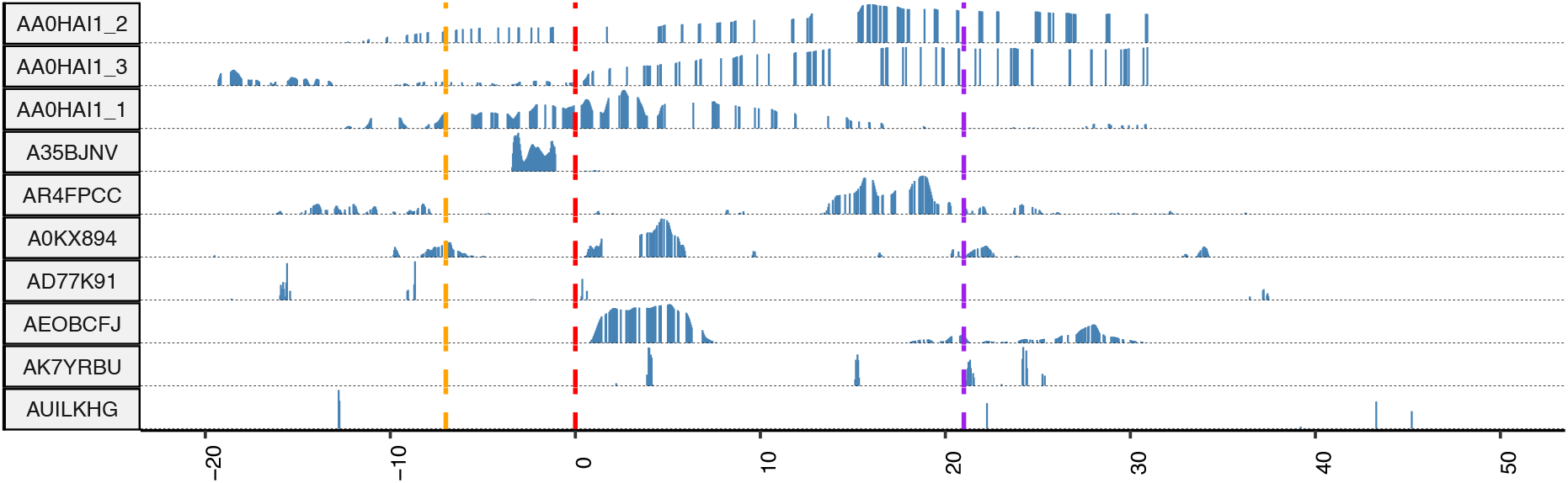
LAAD predictions for the non-COVID-19 group. The x-axis shows days before, during and after the infection and the y axis shows loss values from LAAD (blue bars). The self-reported symptom onset date was shown as a red dotted line, a pre-symptomatic window as a gold dotted line and a post-symptomatic window as a purple dotted line.

**Section 2: LAAD predictions for a healthy group**

**Figure S.2.**
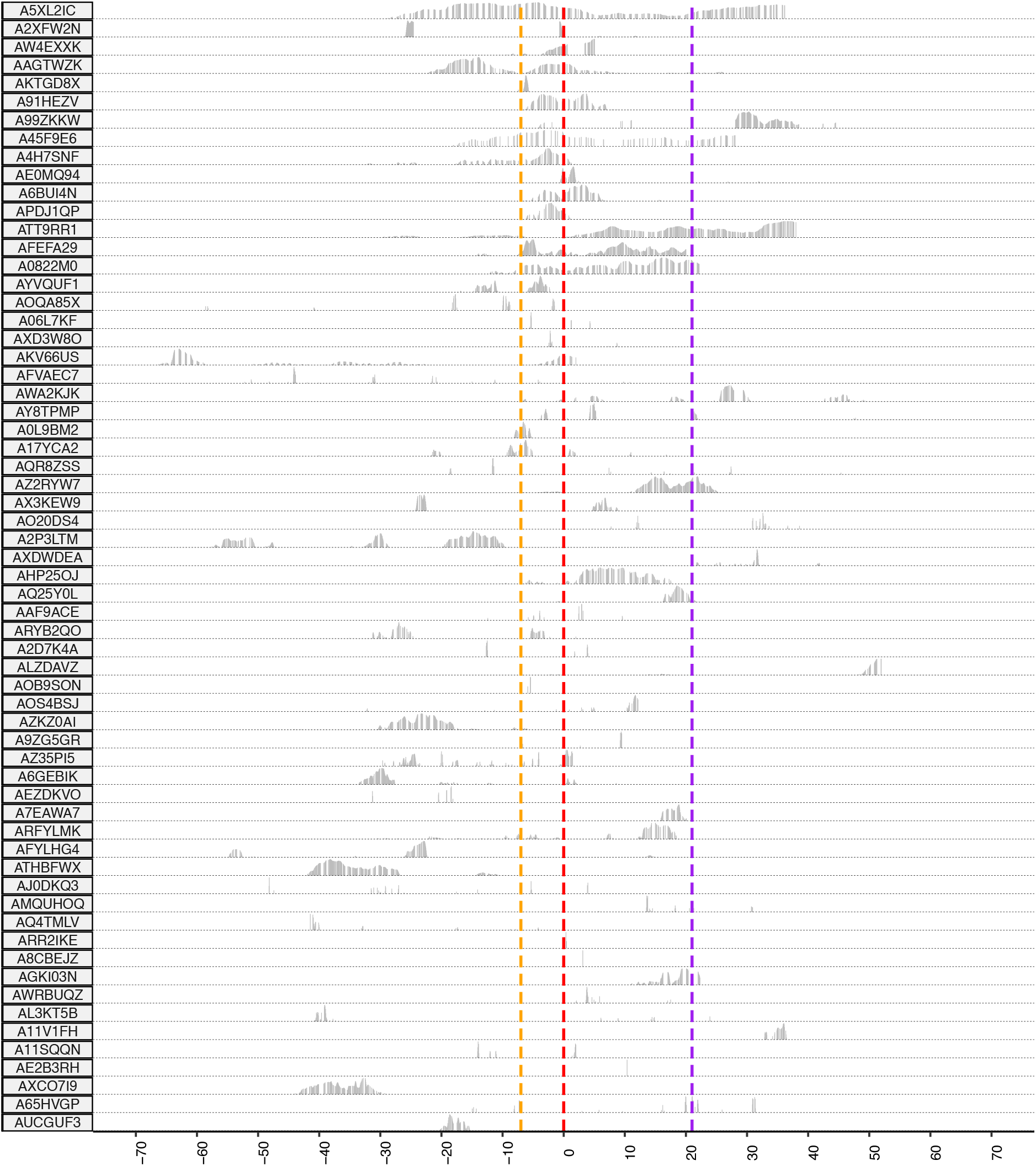
LAAD predictions for a healthy group with randomly chosen symptom onset date. The x-axis shows days before, during and after the infection and the y axis shows loss values from LAAD (blue bars). The self-reported symptom onset date was shown as a red dotted line, a pre-symptomatic window as a gold dotted line and a post-symptomatic window as a purple dotted line.

**Section 3: Number of abnormal RHR hours, and median RHR differences from the baseline in each user during the infectious period**.

**Figure S.3.**
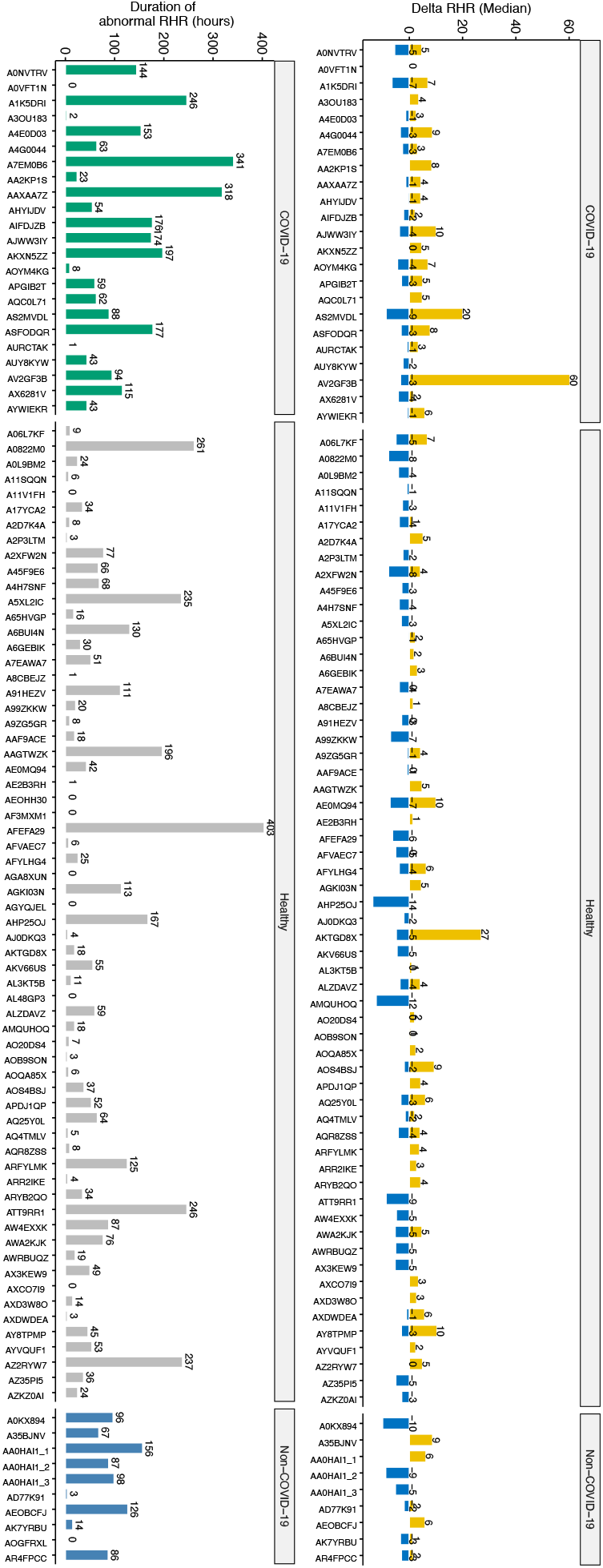
Number of abnormal RHR hours (left) and median RHR differences (right) from the baseline in each user during the infectious period separated by COVID-19, non-COVID-19 and healthy groups. On the left, COVID-19 is shown in green, healthy in grey and non-COVID-19 in blue color. On the right, elevated abnormal RHR during infectious period shown in gold (positive direction) and lowered shown in blue (negative direction).

**Section 4: Training and validation losses in COVID-19+ patients**

**Figure S.4.**
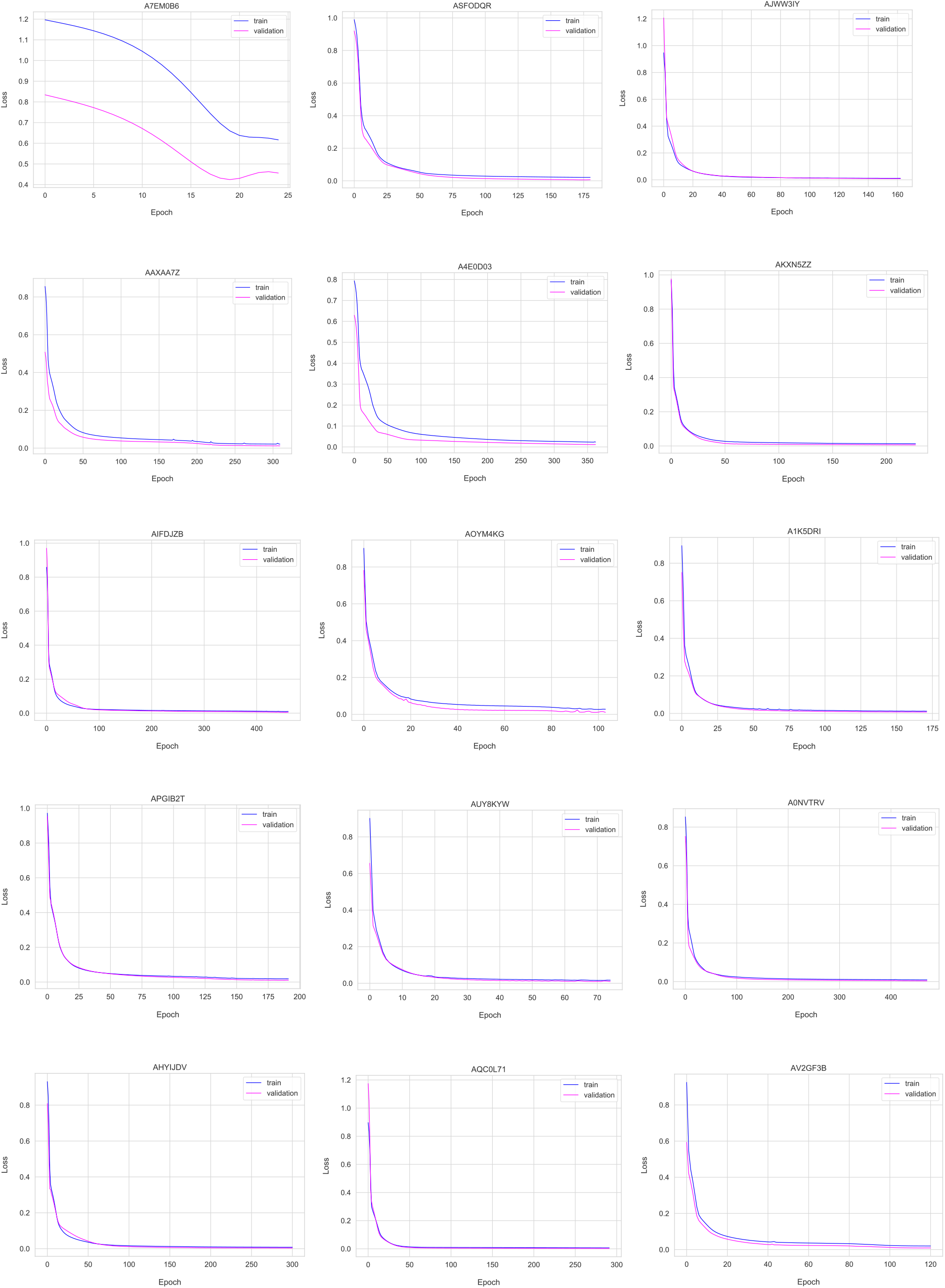
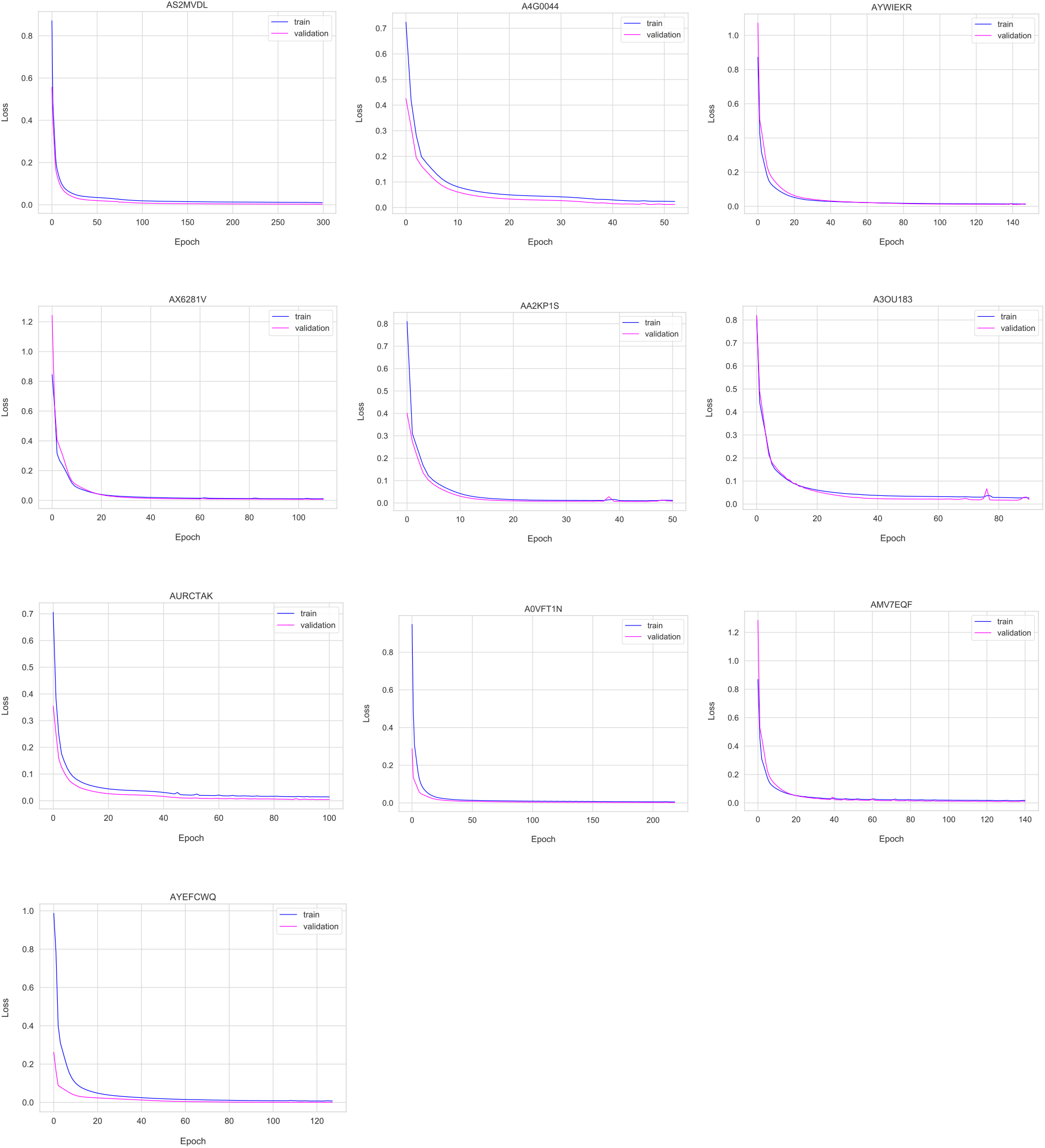
Training (blue) and validation (purple) losses in COVID-19+ patients. Training data were augmented into around eight times the size and used 95% of this augmented data as training and 5% as validation to train the model and plotted reconstruction loss over several epochs using early callbacks. A7EM0B6 had only 4 days of training data before augmentation and showed severe underfitting of the model.

**Section 5: Distributions of training**

**Figure S.5.**
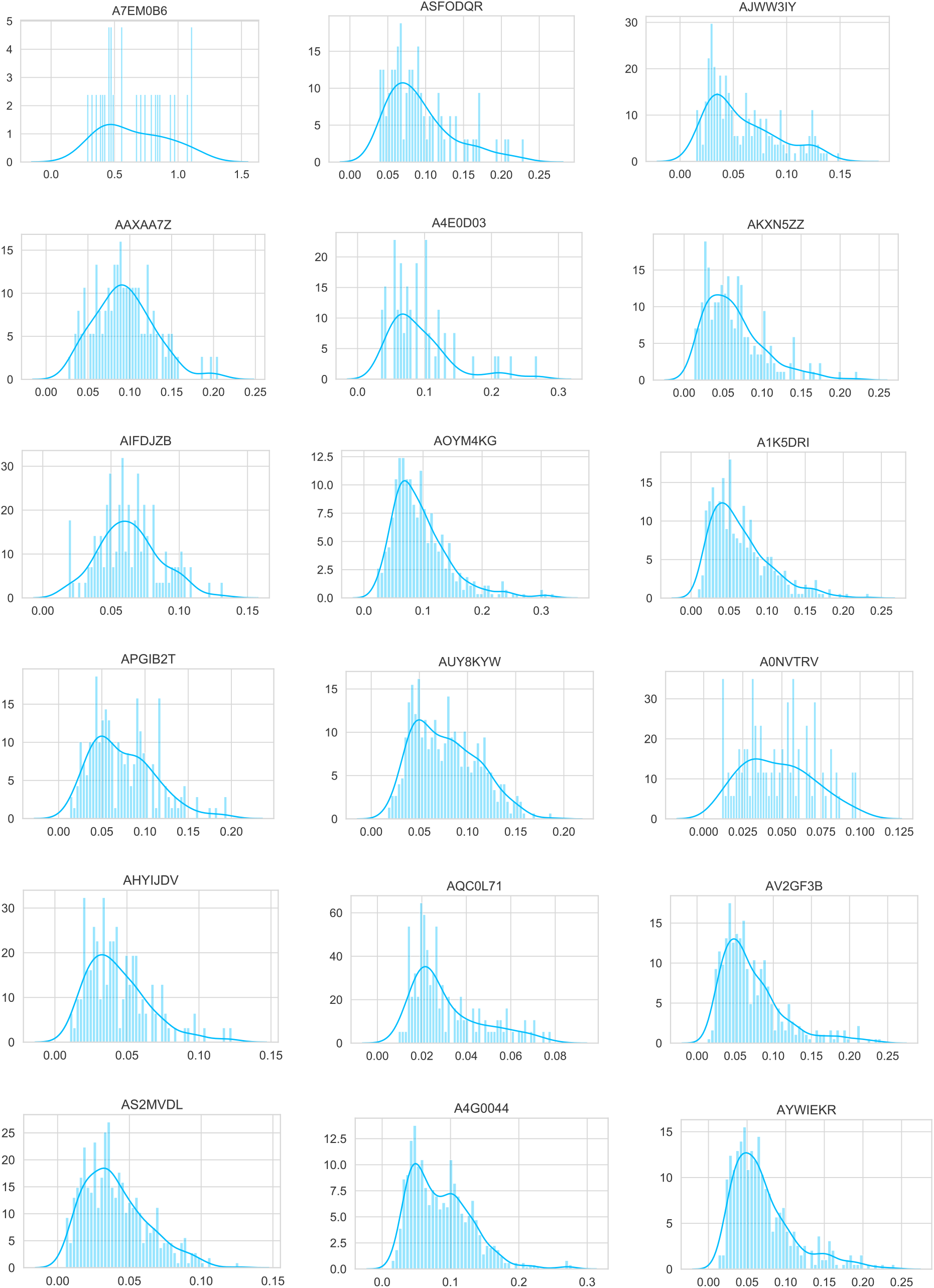
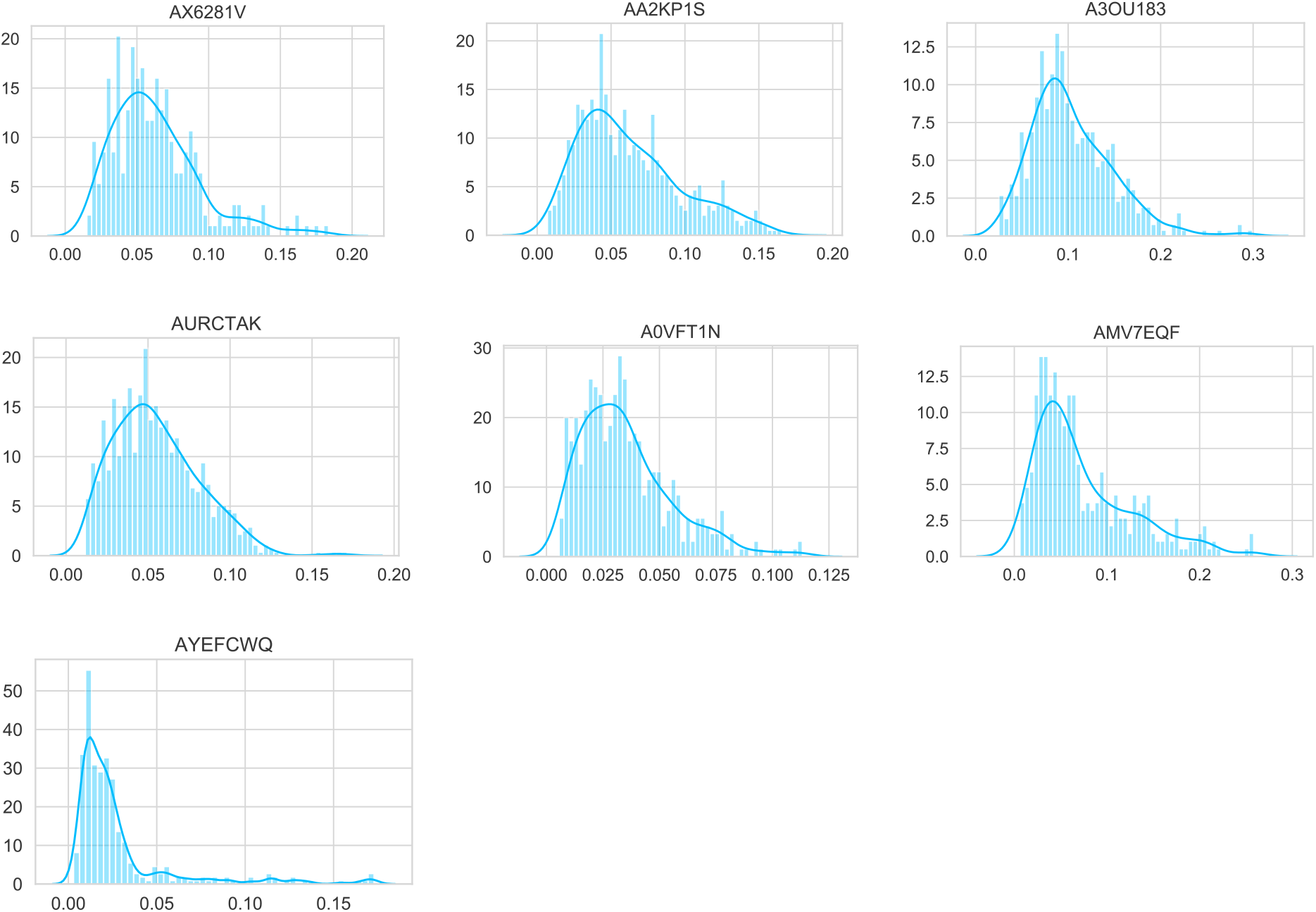
Distributions of training loss (Reconstruction error or Mean Squared Error) used to select the anomaly threshold in COVID-19+ patients.

**Section 6: LAAD predictions in COVID-19+ patients**

**Figure S.6.**
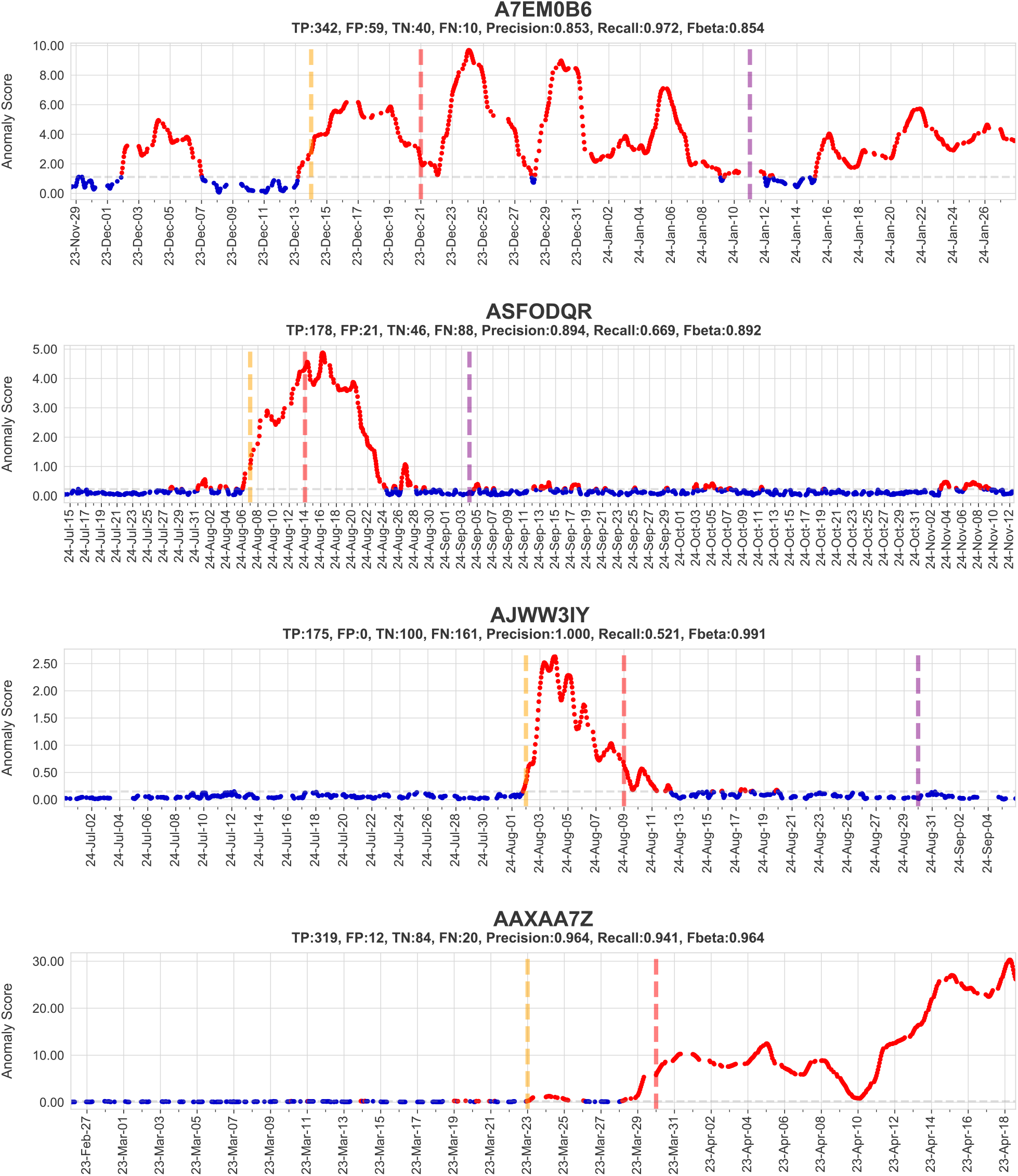
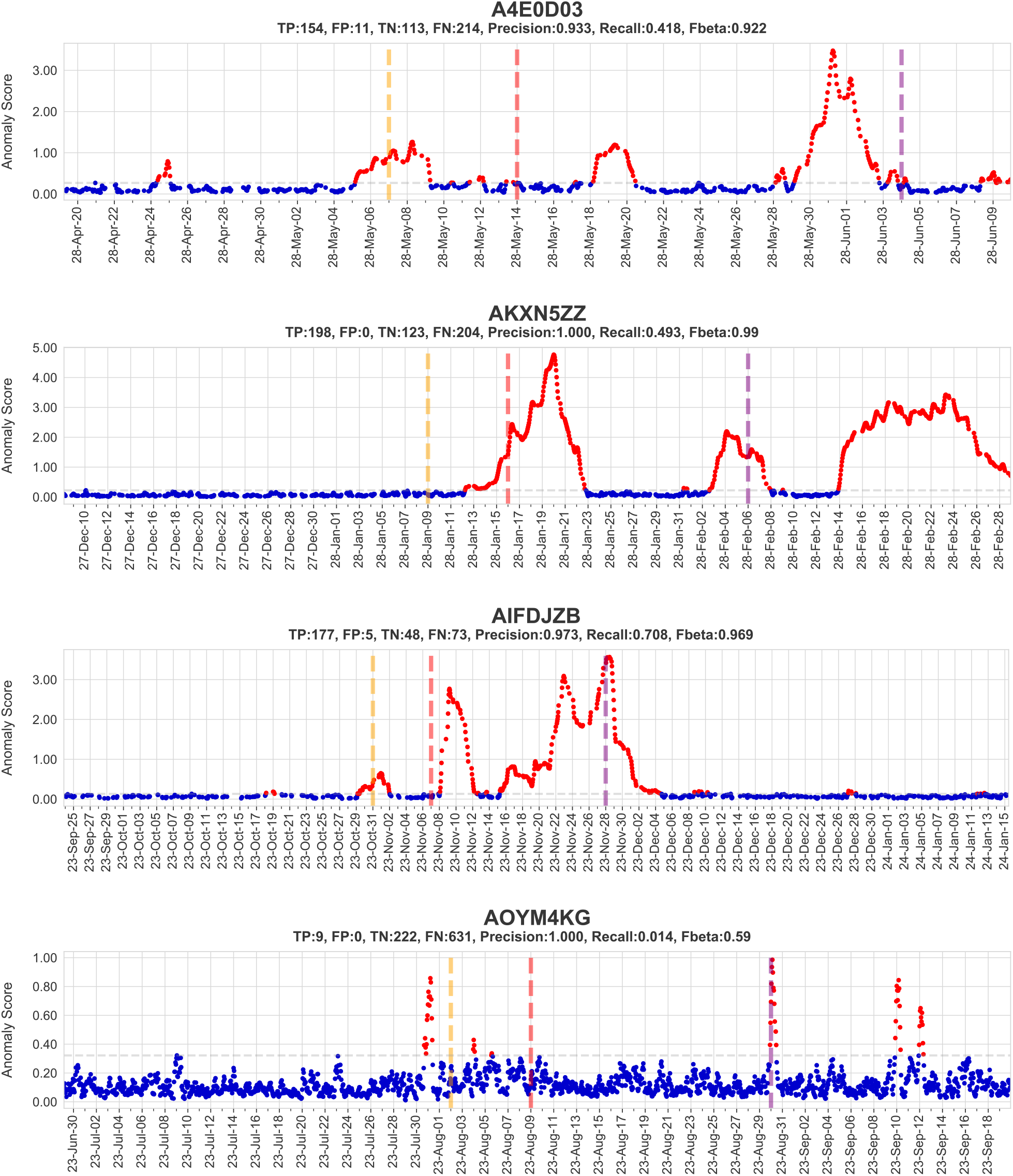
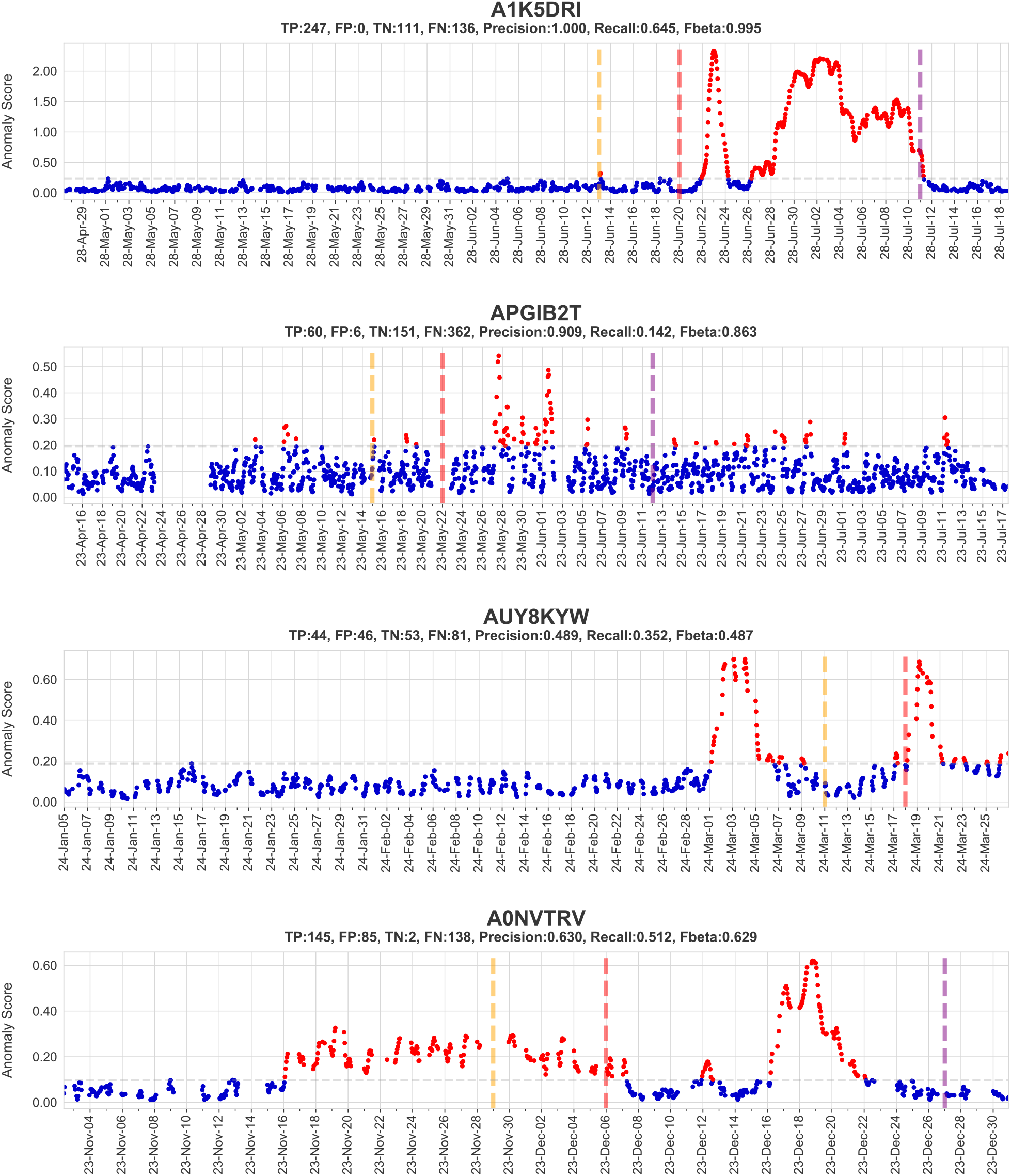
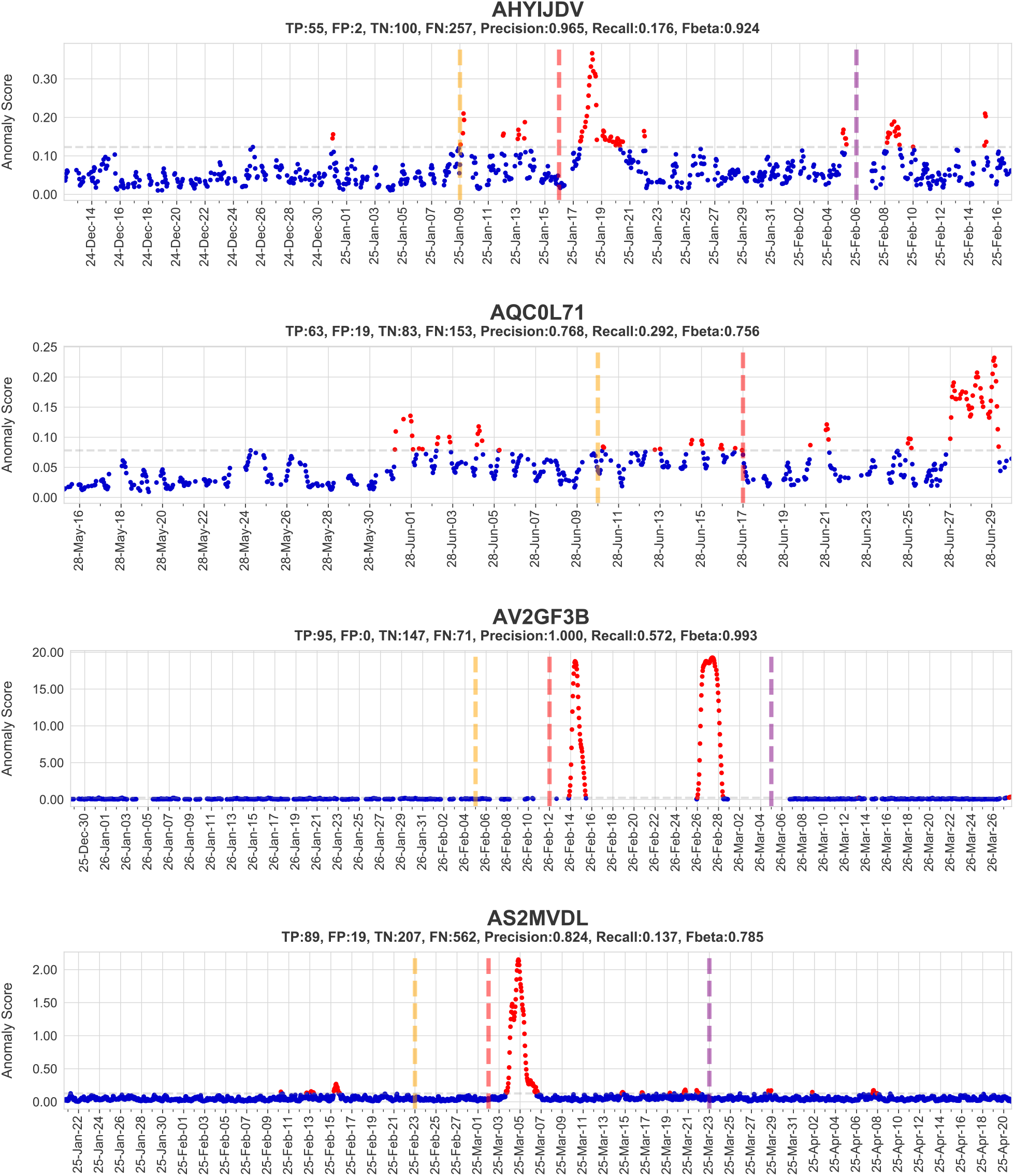
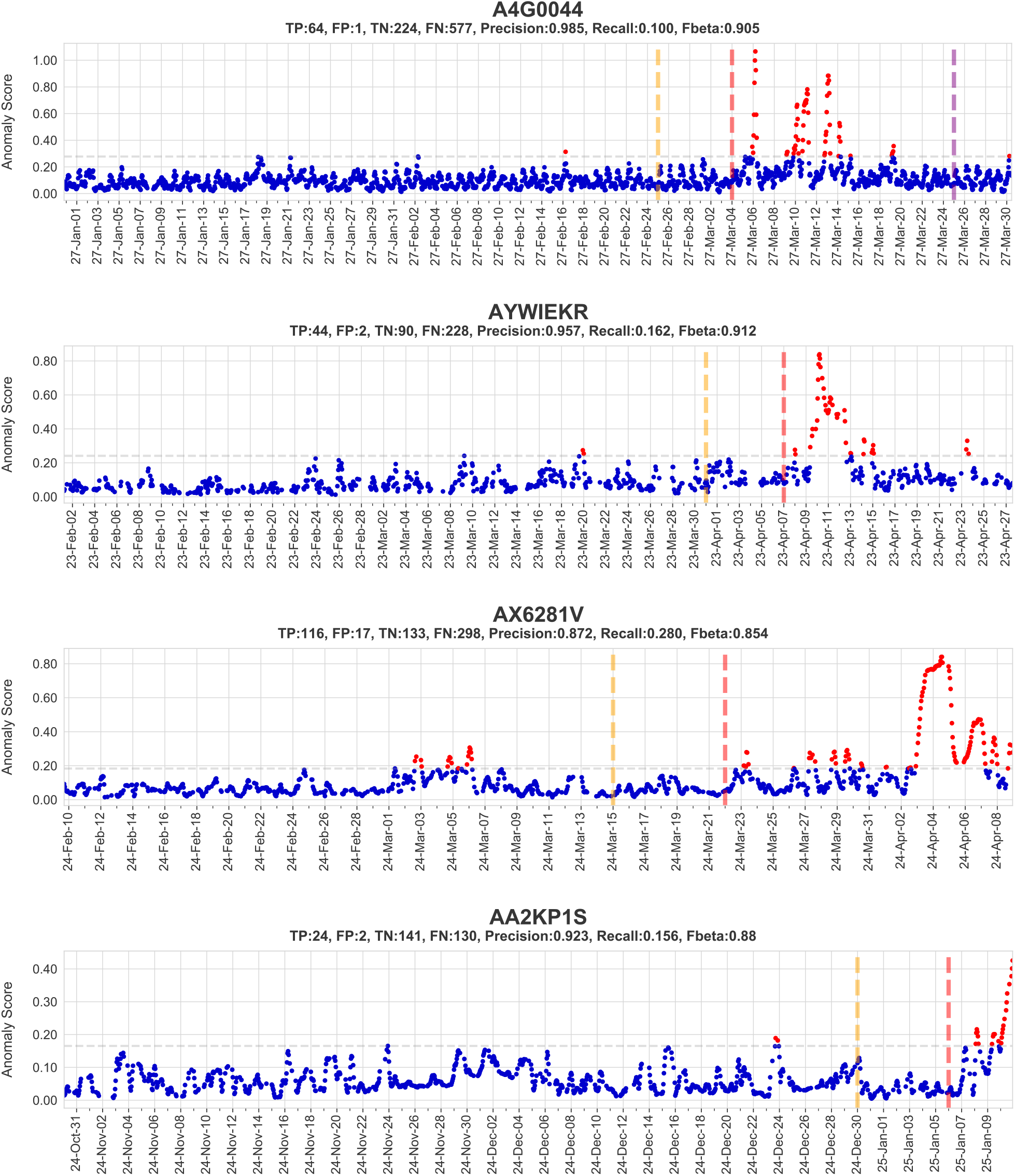
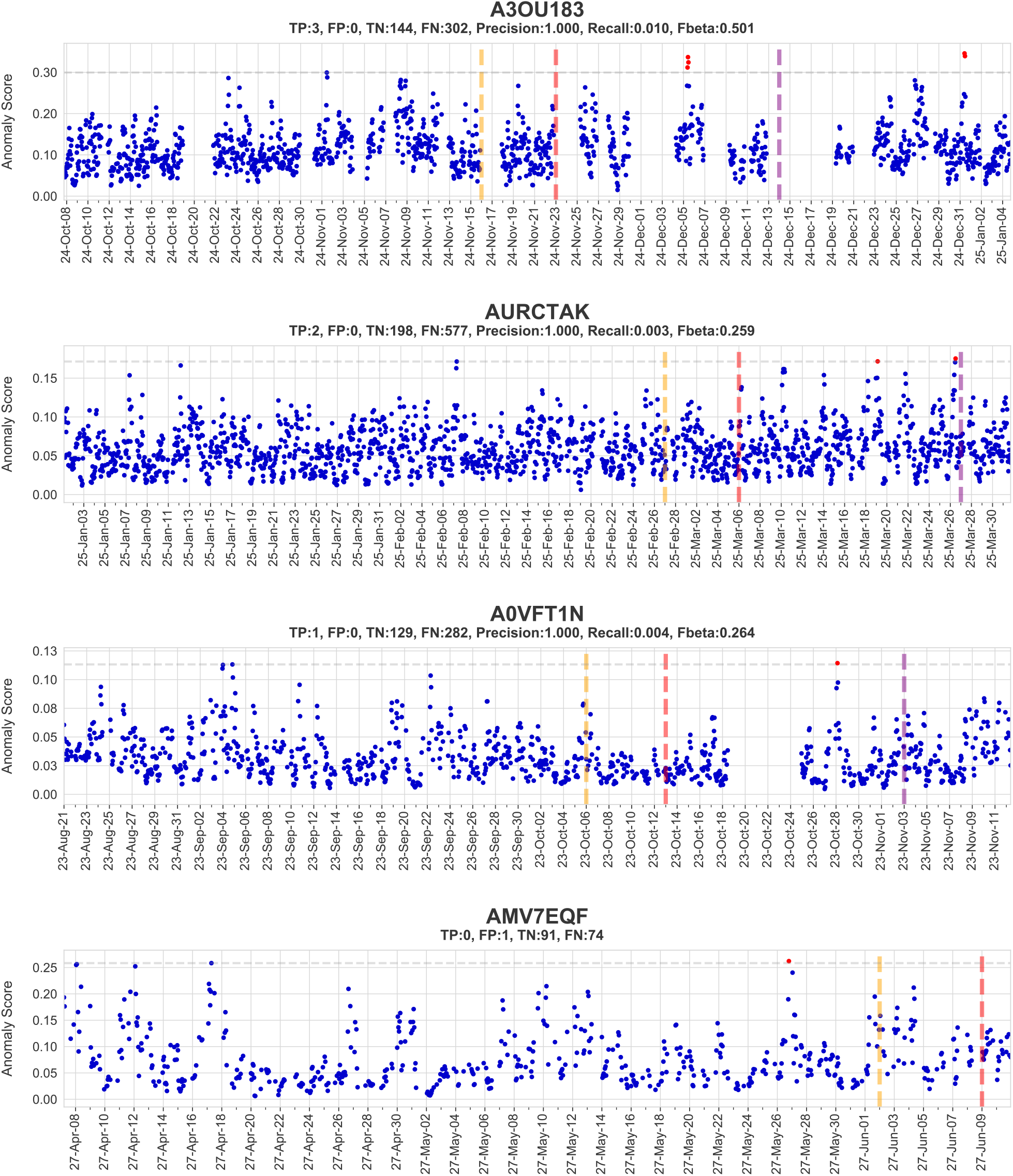
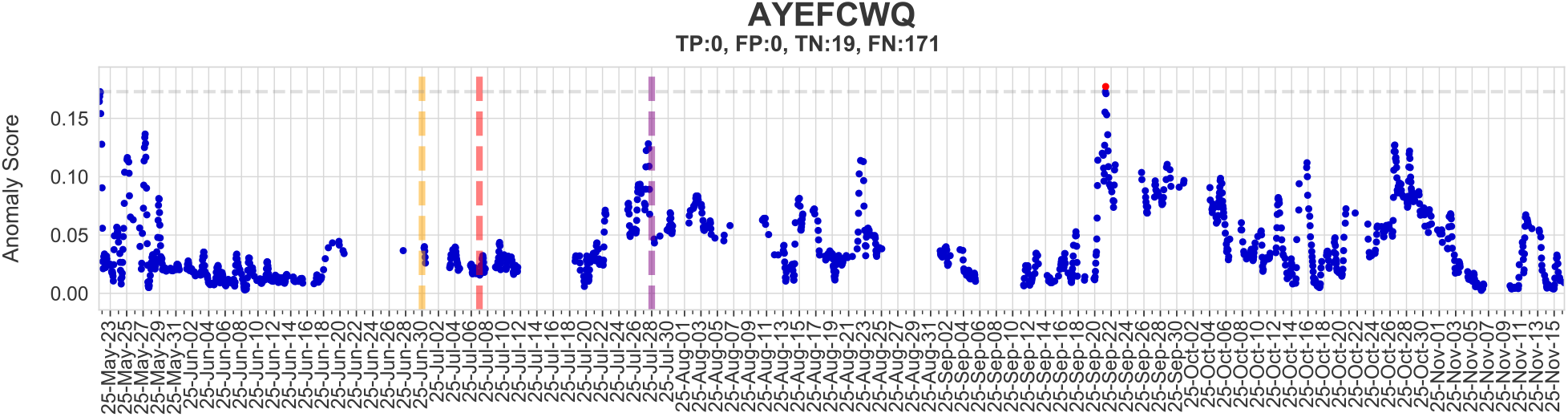
LAAD predictions in COVID-19+ patients (normal – blue, anomaly – red). The red dotted line is the symptom onset date, gold line represents the pre-symptomatic window (7 days before symptom onset) and purple line represents the post-symptomatic window (21 days after symptom onset), and the grey horizontal dotted line represents the selected threshold.

### Supplemental appendix 2

**Section 1 - Supplemental Table 1**. Symptom and diagnosis dates of COVID-19, non-COVID-19 and randomly chosen symptom dates of healthy participants.

**Section 2 - Supplemental Table 2**. Data sizes of COVID-19, non-COVID-19 and healthy participants.

**Section 3 - Supplemental Table 3**. Anomalies predicted by LAAD in COVID-19 patients.

**Section 4 - Supplemental Table 4**. Anomalies predicted by LAAD in non-COVID-19 participants.

**Section 5 - Supplemental Table 5**. Anomalies predicted by LAAD in healthy participants.

**Section 6 - Supplemental Table 6**. COVID-19 patients grouped into early, late depending on time of detection and number of anomalies (signal), strong and weak groups depending on signal strength during the infectious period.

**Section 7 - Supplemental Table 7**. Non-COVID-19 participants grouped into early, late depending on the time of detection and number of anomalies (signal), strong and weak groups depending on signal strength during the infectious period.

**Section 8 - Supplemental Table 8**. Healthy participants grouped into early, late depending on time of detection and number of anomalies (signal), strong and weak groups depending on signal strength during infectious periods.

**Section 9 - Supplemental Table 9**. Evaluation metrics (precision, recall, F-beta score) in COVID-19 patients.

**Section 10 - Supplemental Table 10**. Evaluation metrics (precision, recall, F-beta score) in non-COVID-19 participants.

**Section 11 - Supplemental Table 11**. Evaluation metrics (precision, recall, F-beta score) in healthy participants.

**Section 12 - Supplemental Table 12**. Duration of the abnormal RHR during infectious period in COVID-19 patients, non-COVID-19 and healthy participants. 11 users listed as possible asymptomatic based on more than 89 hours of abnormal RHR during the infectious period.

**Section 13 - Supplemental Table 13**. Delta RHR (the median difference between abnormal RHR from the infectious period and RHR from baseline) in COVID-19 patients, non-COVID-19 and healthy participants. TRUE in the positive column indicates increased RHR and FALSE indicates lowered RHR.

